# Seroprevalence of SARS-CoV-2 infection in the Colombo Municipality region, Sri Lanka

**DOI:** 10.1101/2021.06.18.21259143

**Authors:** Chandima Jeewandara, Dinuka Guruge, Inoka Sepali Abyrathna, Saubhagya Danasekara, Banuri Gunasekera, Pradeep Darshana Pushpakumara, Deshan Madhusanka, Deshni Jayathilaka, Thushali Ranasinghe, Gayasha Somathilaka, Shyrar Tanussiya, Tibutius Tanesh Jayadas, Heshan Kuruppu, Nimasha Thashmi, Michael Harvie, Ruwan Wijayamuni, Lisa Schimanski, T.K. Tan, Pramila Rijal, Julie Xiao, Graham S. Ogg, Alain Townsend, Gathsaurie Neelika Malavige

## Abstract

**Background:** As the Municipality Council area in Colombo (CMC) experienced the highest number of cases until end of January 2021, in Sri Lanka, we carried out a serosurvey prior to initiation of the vaccination program to understand the extent of the SARS-CoV-2 outbreak.

**Methods:** SARS-CoV-2 seropositivity was determined in 2547 individuals between the ages of 10 to 86 years, by the Wantai total antibody ELISA. We also compared to seroprevalence using the haemagglutination test (HAT) to evaluate its usefulness in carrying out serosurveys.

**Results:** The overall seropositivity rate was 24.46%, while seropositivity by HAT was 18.9%. Although the SARS-CoV-2 infection detection rates by PCR were highest in the population between the ages of 20 to 60 years of age, the seropositivity rates were equal among all age groups. The seropositivity rate was highest in the 10 to 20 age group (34.03%), whereas the PCR positivity rates was 9.8%. Differences in the PCR positivity rates and seropositivity rates were also seen in 60- to 70-year-olds (8.9% vs 30.4%) and in individuals >70 year (4.1% vs 1.2%). The seropositivity rates of the females was 29.7% (290/976), which was significantly higher (p<0.002) than in males 21.2% (333/1571).

**Conclusions:** A high seroprevalence rate (24.5%) was seen in all age groups in the CMC suggesting that a high level of transmission was seen during this area. The PCR positivity rates, appear to underestimate the true extent of the outbreak and the age groups which were infected.

## Introduction

Eighteen months following the reporting of the first person infected by the SARS-CoV-2 virus, many countries are currently experiencing the third wave with higher caseloads and mortality rates. The steepest increase in the number of cases globally is seen now, as the outbreak is affecting many countries in South and South East Asia and Latin America [1], which have scarce resources to deal with such large numbers. Similar to the situation in many other South Asian countries, the number of COVID-19 cases is rapidly rising in Sri Lanka. The first patient was detected on the 27^th^ of January 2020, who was a foreign national from China and the first Sri Lankan patient was reported on the 10^th^ of March [2]. Since the detection of the first patient, Sri Lanka went for a strict and extensive lockdown after 10 days (on the 20^th^ of March), which enabled limiting the initial outbreak to certain areas of the Colombo Municipal Council (CMC) area. Although Sri Lanka successfully contained the epidemic until the end of September, with no locally detected cases from August to September 2020, a large outbreak emerged during early October, which rapidly spread island wide. However, as the CMC is the business capital of the country, and also due to extremely overcrowded living conditions, 32,346/89,817 (36.01%) locally detected cases seen by the end of March 2021, were detected within the Colombo district [3]. Of the cases in the Colombo district 14,416 (44.6%) were identified within the CMC.

The CMC has a population of 561,314 individuals living in an area of 37.3 km^2^. The CMC is divided into 6 districts: namely D1, D2A, D2B, D3, D4 and D5 (Figure 1). The overall population density in the whole CMC is 20,187.8 individuals/km^2^, although certain areas have a higher density due to poor-housing conditions and overcrowding. During the second wave in Sri Lanka, which occurred from October 2020 to March 2021, a rapid rise in the number of cases and intense transmission was seen within this area. However, the number of cases were seen to gradually decline from mid-February to end of March 2021, with less than 10 cases per day detected in early April. Administration of the COVID-19 vaccines (Covishield) started on the 29^th^ of January 2021 in Sri Lanka, by initially immunizing health care workers. Immunizing the general public began after the first week of February and due to the high number of cases, the CMC area was prioritized. By mid-March, 20% of the CMC population were vaccinated. Although vaccination itself may have led to a decline in the number of cases in the CMC, it could have also been due to high past infection rates resulting in many individuals being immune to the virus.

**Figure 1:**
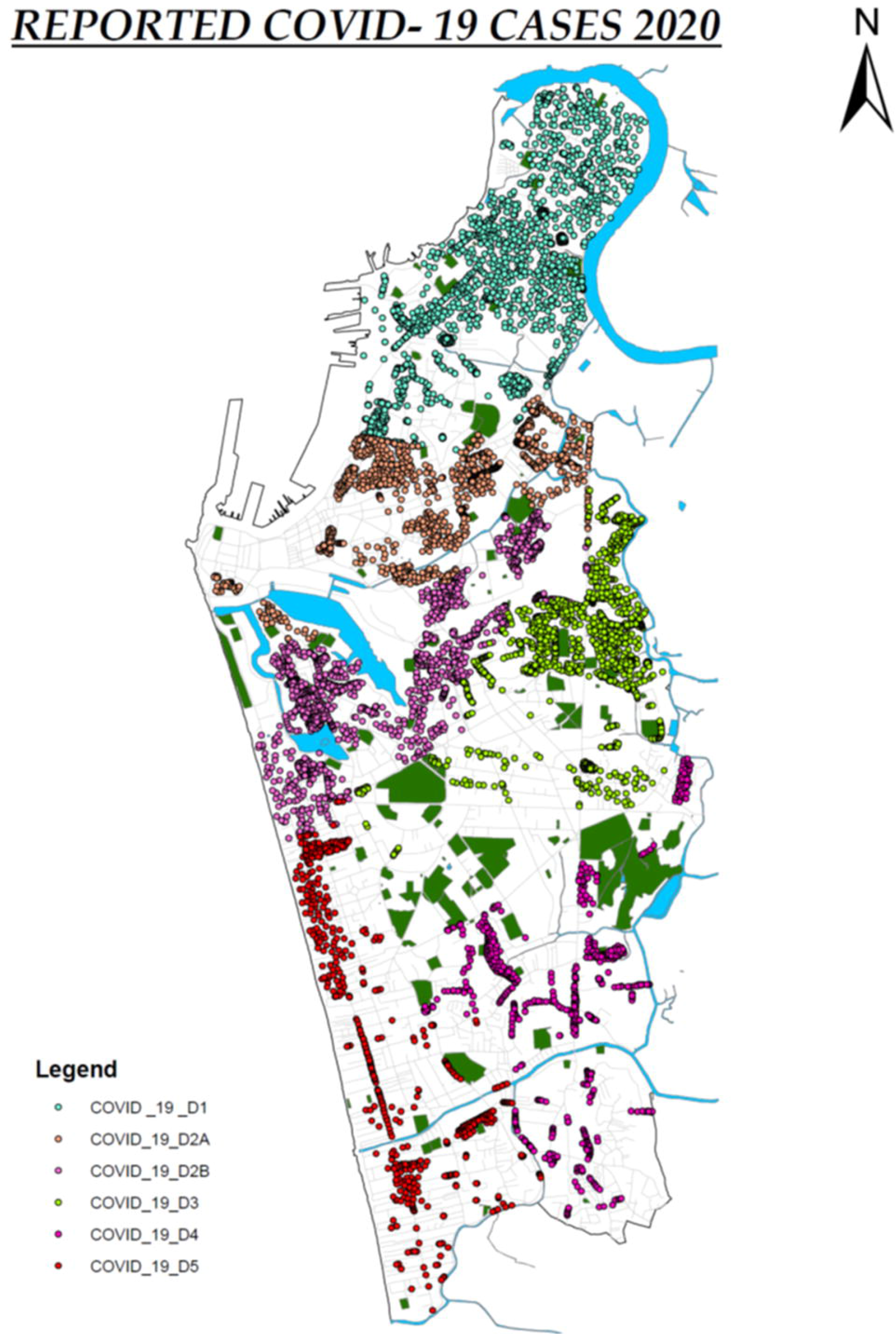
The six districts in the Colombo Municipality Council (CMC) and the locations of cases (identified by qRT-PCR). Each dot denotes an individual person.

In many countries, the reported number of cases do not necessarily reflect the extent of the outbreak, age groups infected and groups at risk, as the majority of infections are asymptomatic and limitations in carrying out quantitative real-time PCR for SARS-CoV2 (qRT-PCR)[4; 5]. It has been estimated that surveillance of SARS-CoV2 with qRT-PCR alone may underestimate the true prevalence by tenfold [6]. For instance, the overall seroprevalence of COVID-19 in India was found to be 7.1% by end of September 2020, which gives infection rates several folds higher than the actually reported number of cases [7]. It is important to carry out serosurvillence studies to understand the true extent of an outbreak in order to understand the future outbreaks that may occur in a particular area and to further understand transmission dynamics and duration of immunity. Therefore, we carried out a serosurveillence study in the CMC at end of January 2021, before the initiation of the COVID-19 vaccination campaign for the general public.

## Methods

### Study population

2547 individuals between the ages of 10 to 86 years were recruited following informed written consent during January 2021 (before administration of COVID-19 vaccines). The population in each of the six districts and the number of individuals from each district recruited for the study is shown in table 1. Blood samples were obtained from the participants at the same time when samples were taken from them for routine PCR testing for SARS-CoV-2. None of the participants had any symptoms at the time of obtaining blood samples and were not previously diagnosed as been infected with the SARS-CoV-2 virus. Basic demographic details such as the age, gender and prior COVID-19 illness were recorded, and blood samples were obtained to determine the seropositivity status. Ethics approval was obtained from the Ethics Review Committee of the University of Sri Jayewardenepura.

**Table 1:** The number of individuals in each of the CMC districts and the proportion samples from each district.

| District number | Total population of the district | Number recruited to the study (%) | PCR positivity (%) | No of PCR+ cases/100,000 population |
| --- | --- | --- | --- | --- |
| D1 | 131012 | 170<br>(0.13%) | 3663<br>(2.79%) | 2795.92 |
| D2A | 137644 | 710<br>(0.52%) | 3290<br>(2.39%) | 2390.22 |
| D2B | 61312 | 477<br>(0.78%) | 2676<br>(4.36%) | 4364.56 |
| D3 | 89855 | 466<br>(0.52%) | 2571<br>(2.86%) | 2861.28 |
| D4 | 80839 | 210<br>(0.26%) | 1282<br>(1.59%) | 1585.87 |
| D5 | 60652 | 514<br>(0.85%) | 934<br>(1.54%) | 1539.93 |
| Total | 561314 | 2547<br>(0.45%) | 14416 | 2568.26 |

### Detection of total antibodies to SARS-CoV-2

SARS-COV-2 specific total antibody (IgM, IgG and IgA) responses were assessed using WANTAI SARS-CoV-2 Ab ELISA (Beijing Wantai Biological Pharmacy Enterprise, China). This assay is specific for the Receptor Binding Domain (RBD) and was shown to have a sensitivity of 98% [8] and was found to be 100% specific based also on control serum samples obtained in 2018, in Sri Lankan individuals. The assay was carried out and results were interpreted according to manufacturers’ instructions.

### Haemagglutination test (HAT) to detect antibodies to the receptor binding domain (RBD)

The HAT assay is very cheap tool, that does not require any specific equipment that detects antibodies to the RBD. In order to compare the usefulness of the HAT assay in comparison to the commercially available Wantai total antibody assay in determining serosurveys, we used the HAT assay in a subset of individuals (n=1413). The HAT was carried out in a subset of these individuals (n=1413) as previously described [9] using method (1). The HAT assay detects haemagglutination of red cells labelled with the IH4-RBD reagent. IH4-RBD is a nanobody against a conserved glycophorin A epitope on red cells, linked to the RBD of SARS-CoV-2. Any antibody in the test serum specific for the RBD that can cross-link and agglutinate the red cells will be detected. The HAT was shown to have a sensitivity ∼90% and specificity >99% several weeks after a PCR diagnosed symptomatic infection^9^. Briefly, red blood cells from an O negative donor diluted in PBS 1:20 (∼2% Red Cells) were mixed with 50ul of the IH4-RBD reagent (2ug/ml stock) and 50ul of 1:20 Serum (2.5ul serum in the reaction well) and incubated for one hour at room temperature to give a dilution of 1:40. Phosphate buffered saline was used as a negative control in place of the diluted serum. At the end of the incubation the plate was tilted for 20 seconds and then photographed. The photograph of the plate was read by two independent readers to examine the “teardrop” formation indicative of a negative result. A complete absence of “teardrop” formation was scored as positive, and any flow of “teardrop” was scored as negative. We have confirmed that this assay is negative in >99% of individuals prior to infection with SARS-CoV-2 (manuscript under review).

## Results

The mean age of the study population was 43.6 (SD±16.07) and 1571 (61.7%) were males. The overall seropositivity as measured by the Wantai total antibody assay was 24.46% (623/2547). The seropositivity rates of the females was 29.7% (290/976), which was significantly higher (p<0.002) than in males 21.2% (333/1571). We also used the haemagglutination test (HAT) to measure antibodies to the RBD where the RBD of the virus is linked to a nanobody IH4, specific for a conserved epitope within glycophorin A on red blood cells (RBCs)[9]. A HAT titre of 1:40 was considered as positive for the presence of RBD-specific antibodies. The overall seropositivity rate by the HAT was 18.9% (267/1413). The age stratified seroprevalence for the total SARS-CoV2 specific antibodies and the HAT assay is shown in table 2. There was no significant correlation of the total antibody positivity and HAT positivity (Spearmans R=0.35, p=0.44).

**Table 2:** Age stratified seroprevalence rates for SARS-CoV2 in different age groups.

| Age group | Seropositive rates by<br>Wantai<br>N=2547 | Seropositive rates by<br>HAT<br>N=1413 | PCR positivity rates<br>N=14,416 |
| --- | --- | --- | --- |
| 10 – 20 | 65/191<br>(34.03%) | 36/126<br>(28.57%) | 1418<br>(9.84%) |
| 21 - 30 | 83/448<br>(18.53%) | 31/253<br>(12.25%) | 2864<br>(19.87%) |
| 31 - 40 | 104/461<br>(22.56%) | 47/253<br>(18.58%) | 2893<br>(20.07%) |
| 41 - 50 | 125/484<br>(25.83%) | 45/265<br>(16.98%) | 2801<br>(19.43%) |
| 51 - 60 | 129/507<br>(25.44%) | 54/273<br>(19.78%) | 2556<br>(17.73%) |
| 61 - 70 | 89/293<br>(30.38%) | 39/179<br>(21.79%) | 1294<br>(8.98%) |
| Over 70 | 28/163<br>(17.18%) | 15/64<br>(23.48%) | 590<br>(4.09%) |
| Total | 623/2547 | 267/1413 | 14416 |

There was no significant difference between age and seropositivity for either assay, with the seropositivity rates been similar in all age groups (Figure 2A). However, the seropositivity rates were slightly higher in the 10 to 20 age group from both the Wantai total antibody assay (34.03%) and HAT (28.57%), compared to other age groups, although this was not statistically significant.

**Figure 2:**
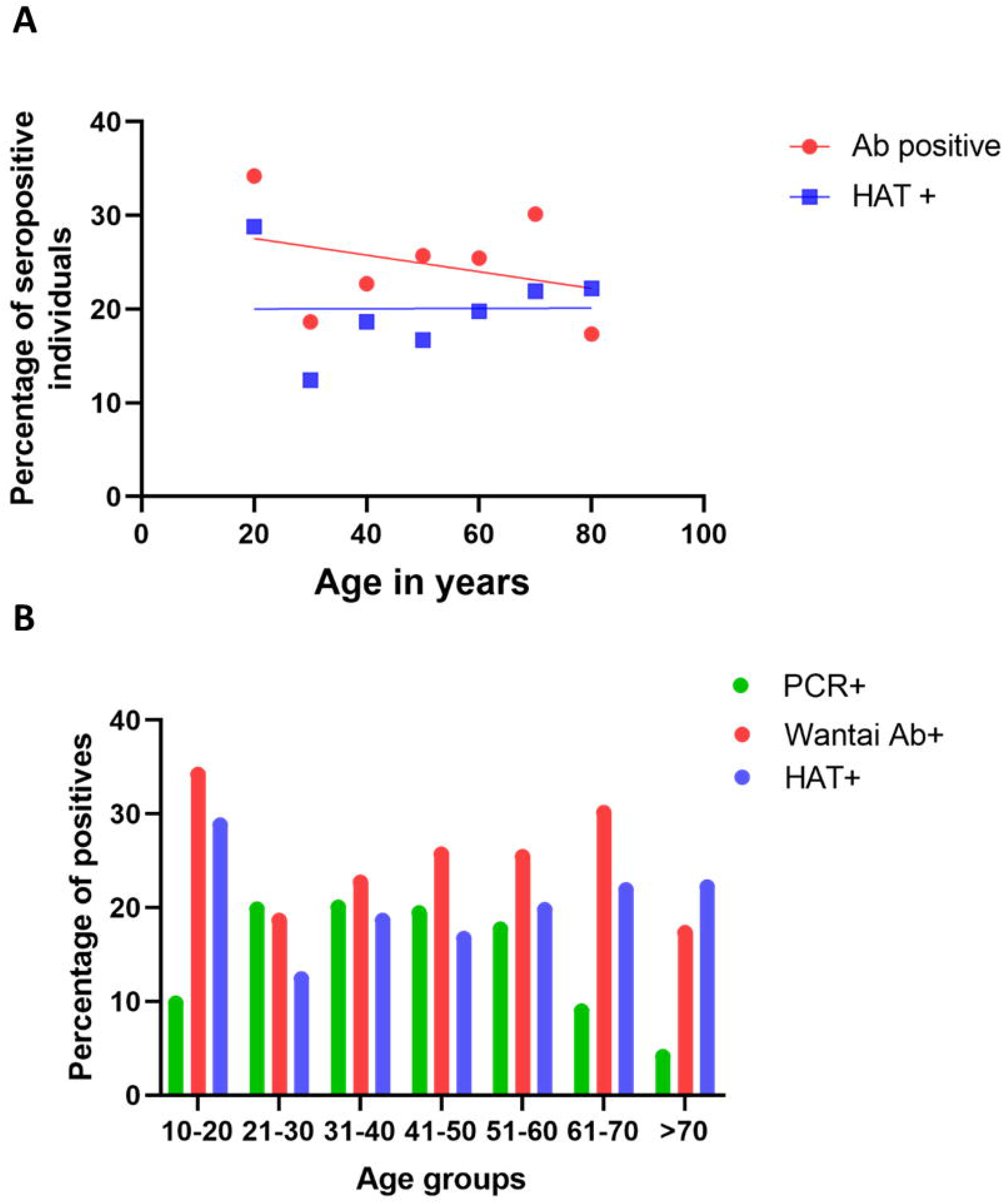
Seropositivity rates and infection rates in different age groups. The seropositivity rates were assessed in each age group by the Wantai total antibody assay (Wantai Ab+) and the HAT assay (HAT+), and the positivity rates of each assay was correlated with the age (A). The PCR positivity rates in each age group and the Wantai (Ab+) and HAT (HAT+) positivity rates were compared in each age group (B).

The Wantai total antibody assay measures the presence of SARS-CoV2 specific IgM, IgG and IgA antibodies to the RBD, while the HAT assay measures any antibodies to the RBD that can cross-link and agglutinate red cells. The total level of antibody to RBD detected in ELISA is known to correlate with viral neutralising titre[10], and we have confirmed this for the titre detected in the HAT assay (manuscript in preparation). Although a higher number of individuals were positive to the Wantai assay in individuals <70 years of age, in those who were >70 years, the HAT positivity rates were higher (23.48%) compared to the Wantai assay, although this was not significant (p=0.27). In those who were <30 years of age and between the ages of 30 to 60, 0.53% and 0.76% were seropositive by HAT, but negative by Wantai. In the >60-year-old age group, 1.65% who were negative by Wantai were positive by HAT, which was significantly higher (p=0.002) when compared to younger age groups. The HAT assay has been shown to have a higher sensitivity early after infection [9], so these differences may reflect the timing of infection prior to recruitment to the study.

The Wantai and HAT seropositivity rates differed between females and males. While 24.95% of females were positive for SARS-CoV-2 by both the Wantai and HAT assays, the positivity rate of males was 13.76%, which was significantly lower (p < 0.002). 1.1% of females were only positive by HAT (negative by Wantai), whereas 0.69% of men were only positive by HAT.

### qPCR positivity in different age groups vs seropositivity

As a large number of cases have been detected in the CMC during this time period, qRT-PCRs were carried out on all primary contacts and most of the secondary contacts. Of the total PCR positives (n=14,416), 11,108 (77.1%) were between the ages of 20 to 60 years. While the PCR positivity rates (infection detection rates) were between 17.7% to 20.05% in the age groups from 20 to 60 years, it was 9.8% in 10- to 20-year-old individuals and 8.9% in 60- to 70-year-olds (Figure 2B). Although infection detection rates were less in those <20 years and those >60 years, the seropositivity rates were higher in the 10- to 20-year-old age group (Figure 2B).

### Seropositivity rates in different districts of the CMC

As shown in table 1, the prevalance rates varied in different districts, with district D2B and D3 reporting a very high number of cases/100,000 population identified through PCR compared to other districts. In order to determine if the qRT-PCR positivity rates indicate the true extent of the outbreaks in these different districts, we assessed the differences in the seropositivity rates in these six districts. Interestingly, although the infection rates were highest in D1 (25.4%), the seropositivity rates were lowest in D1 (4%). In contrast, D2A which had a PCR+ rate of 22.8% had a seropositivity of 31% and D3, which had a PCR+ rate of 17.8% had a seropositivity rate of 29.5% (table 3).

## Discussion

In this study, we have carried out a serosurveillance for COVID-19 in the CMC area, which experienced the highest number of cases (16.1%) in the whole country. Our data showed that the overall seropositivity rate was 24.46% in this area, until the end of January 2021. This is several fold higher than the prevalence of COVID-19 based on PCR positivity, which is 2,568/100,000 population. Based on the seropositivity rates of 24.46%, 138,276 individuals are likely to have been infected with the SARS-CoV-2 virus compared to the reported PCR positive cases of 14,416. Therefore, infection detection rates by PCR, appeared to have underestimated the actual number of infections by 9.59-fold. This is comparable to studies carried out elsewhere, which have shown that the serologically detected cases out number the virologically confirmed SARS-CoV-2 infection by 10-fold [6]. In this study we evaluated the usefulness of the HAT to determine seroprevalence, which showed a seropositivity rate of 18.9%. Although the seropositivity rates from HAT were slightly lower than with the Wantai total antibody assay, our data show that the HAT assay appears to be a sensitive tool, that can be used to carry out serosurveys in resource poor settings as it is a cheap assay that does not require any equipment.

Although the overall seroprevalence was 24.46%, certain districts in the CMC (D2A, D2B and D3) had higher seroprevalence rates (26.2% to 39%) compared to D4 which only had a seroprevalence rate of 3.33%. In D1, although the seroprevalence was 14.76%, certain areas in this district had a very high infection rate and were isolated very early, and therefore, it would have curtailed the spread to the rest of the D1 district. These overall differences between the districts reflect the population density and the housing conditions in these districts, with the districts with high seroprevalence having more overcrowded areas, with poor housing conditions. Such similar differences have been observed in many states in India, where the slum areas reported seroprevalence rates between 52.6% to 58.7% compared to 12 to 17.9% in non-slum areas [11]. Although the overall seroprevalence rates in the CMC was less than urban areas in India, it was higher than many areas in Europe (Spain, Sweden, Switzerland and Germany), which reported a seroprevalence between 5% to 13.6% and Iran (22.16%), which reported higher infection rates [12; 13; 14; 15]. Although these seroprevalence studies in other countries were carried out during 2020, Sri Lanka did not experience any large-scale outbreaks of COVID-19 until October 2020, and a serosurvey carried out in May 2020, in an area in the CMC where the first outbreak was seen, showed a seroprevalence rates of 1.5% in individuals living in that area (manuscript under review). Therefore, the large outbreak that was seen mainly in the CMC area, which began in early October 2020, appeared to have rapidly spread, resulting in 24.46% of individuals being infected within four months.

Although the SARS-CoV-2 infection detection rates were highest in the population between the ages of 20 to 60 years of age (working population), the seropositivity rates were equal among all age groups. For instance, although the PCR positivity rates were 9.8% in 10- to 20-year-old 8.9% in 60- to 70-year-olds, and 4.1% in individuals >70 years, their seropositivity rates were 36.8%, 32.1% and 26.6% respectively. Therefore, it is likely that individuals of all age groups were equally infected by the SARS-CoV-2 virus, although the infections were only detected by PCR in working age groups, possibly because more PCRs were carried out in the working population. However, interestingly, the seroprevalence rates were significantly higher in females (29.7%) when compared to males (21.1%). Although the reasons for these differences are not clear, it is possible that more females were exposed to other infected individuals, while carrying out their daily activities using common water and washroom facilities, available in these overcrowded housing settings. Interestingly, significantly more females (24.9%) were positive for SARS-CoV-2 by both the Wantai and HAT assays, while positivity rates of males were only 13.76%. The HAT assay detected any agglutinating antibodies to the RBD of the spike protein while the Wantai total antibody assay detect IgG, IgM and IgA antibodies to the RBD.

In summary, we assessed the seroprevalence in the CMC area in Colombo, which experienced the highest number of cases from October 2020 to January 2021. Our data show that the serologically detected infections outnumbered the PCR detected cases by almost 10-fold, which demonstrates the importance of seroprevalence studies in identifying the true extent of an outbreak.

## Data Availability

All data is available within the manuscript file and the figures.

